# Favipiravir, lopinavir-ritonavir or combination therapy (FLARE): a randomised, double blind, 2x2 factorial placebo-controlled trial of early antiviral therapy in COVID-19

**DOI:** 10.1101/2022.02.11.22270775

**Authors:** David M Lowe, Li-An K Brown, Kashfia Chowdhury, Stephanie Davey, Philip Yee, Felicia Ikeji, Amalia Ndoutoumou, Divya Shah, Alexander Lennon, Abhulya Rai, Akosua A Agyeman, Anna Checkley, Nicola Longley, Hakim-Moulay Dehbi, Nick Freemantle, Judith Breuer, Joseph F Standing, FLARE Investigators

## Abstract

**Background:** Early antiviral treatment is effective for COVID-19 but currently available agents are expensive. Favipiravir is routinely used in many countries, but efficacy is unproven. Antiviral combinations have not been systematically studied. We aimed to evaluate the effect of favipiravir, lopinavir-ritonavir or the combination of both agents on SARS-CoV-2 viral load trajectory when administered early.

**Methods:** We conducted a Phase 2, proof of principle, randomised, placebo-controlled, 2×2 factorial, double-blind trial of outpatients with early COVID-19 (within 7 days of symptom onset) at two sites in the United Kingdom. Participants were randomised using a centralised online process to receive: favipiravir (1800mg twice daily on Day 1 followed by 400mg four times daily on Days 2-7) plus lopinavir-ritonavir (400mg/100mg twice daily on Day 1, followed by 200mg/50mg four times daily on Days 2-7); favipiravir plus lopinavir-ritonavir placebo; lopinavir-ritonavir plus favipiravir placebo; or both placebos. The primary outcome was SARS-CoV-2 viral load at Day 5, accounting for baseline viral load. ClinicalTrials·gov: NCT04499677.

**Findings:** Between 6 October 2020 and 4 November 2021, we recruited 240 participants. For the favipiravir+lopinavir-ritonavir, favipiravir+placebo, lopinavir-ritonavir+placebo and placebo-only arms, we recruited 61, 59, 60 and 60 participants and analysed 55, 56, 55 and 58 participants respectively who provided viral load measures at Day 1 and Day 5. In the primary analysis, the mean viral load in the favipiravir+placebo arm had decreased by 0.57 log_10_ (95% CI -1.21 to 0.07, p=0.08) and in the lopinavir-ritonavir+placebo arm by 0.18 log_10_ (95% CI -0.82 to 0.46, p=0.58) more than in the placebo arm at Day 5. There was no significant interaction between favipiravir and lopinavir-ritonavir (interaction coefficient term: 0.59 log_10_, 95% CI -0.32 to 1.50, p=0.20). More participants had undetectable virus at Day 5 in the favipiravir+placebo arm compared to placebo only (46.3% vs 26.9%, odds ratio (OR): 2.47, 95% CI 1.08 to 5.65; p=0.03). Adverse events were observed more frequently with lopinavir-ritonavir, mainly gastrointestinal disturbance. Favipiravir drug levels were lower in the combination arm than the favipiravir monotherapy arm.

**Interpretation:** At the current doses, no treatment significantly reduced viral load in the primary analysis. Favipiravir requires further evaluation with consideration of dose escalation. Lopinavir-ritonavir administration was associated with lower plasma favipiravir concentrations.

**Funding:** LifeArc, UK.

## Introduction

Severe acute respiratory syndrome coronavirus 2 (SARS-CoV-2) continues to represent a major threat to global health. Interrupting viral replication in early infection reduces the risk of COVID-19 disease progression and hospitalisation [1–4], and this is the most logical time to employ antiviral medications. Efficacy has been demonstrated for neutralising monoclonal antibody treatments, but these are vulnerable to loss of potency with new viral variants as observed with the B.1.1.529 (omicron) variant [5]. Furthermore, the cost of available oral antiviral and monoclonal treatments is prohibitive for many countries.

A general principle of antiviral chemotherapy is that multiple agents with different modes of action are often required, which can be particularly pertinent in the case of repurposed drugs where antiviral potency using monotherapy may be limited. Combination therapy using a polymerase inhibitor combined with a protease inhibitor, thereby targeting sequential steps in the viral replication pathway, is a potential strategy [6]. Where SARS-CoV-1 was treated with the polymerase inhibitor ribavirin in combination with the protease inhibitor lopinavir-ritonavir, and when this combination was initiated immediately upon diagnosis, a significant decrease in mortality was seen compared with historical controls [7]. Another study of this combination showed reduced mortality and need for intubation when therapy was given early, but late rescue treatment had no effect [8]. Early post-exposure prophylaxis against Middle East Respiratory Syndrome (MERS-CoV) in healthcare workers also showed that ribavirin plus lopinavir-ritonavir reduced the incidence of infection from 28% to 0% [9].

In early 2020 it was shown that whilst ribavirin had little effect on SARS-CoV-2 viral replication *in vitro*, the orally available polymerase inhibitor favipiravir did have an *in vitro* potency within clinically achievable range [10] [Supplementary Figure 1]. Whilst subsequent *in vitro* results have been less promising, high-dose favipiravir achieving concentrations commensurate with human exposures reduced viral load and lung histopathology in hamsters [11]. Early observational clinical studies reported benefits of favipiravir in COVID-19 patients [12, 13]. Favipiravir generic formulations are now in widespread use for COVID-19 in some regions of the world, but high-quality evidence on its effect in early treatment is lacking. A recent pre-print suggested that favipiravir (as monotherapy and taken with a twice daily dosing regimen) did not impact time to viral clearance [14].

Whilst the HIV protease inhibitors tipranavir and nelfinavir showed higher *in vitro* potency against SARS-CoV-2 than lopinavir-ritonavir [10] safety concerns and limited clinical experience with both agents meant that we chose to study lopinavir-ritonavir. Both lopinavir and ritonavir, which is used as a pharmacokinetic booster to lopinavir, have modest anti-SARS-CoV-2 activity *in vitro* [10] which was predicted to yield around up to 30% inhibition of viral replication at the licensed dose [Supplementary figure 1]. In line with this, lopinavir-ritonavir monotherapy did not improve clinical outcomes in platform trials on hospitalised patients [15, 16]. However, viral dynamic modelling suggests that drugs with lower potency may nevertheless inhibit viral replication if started earlier [17, 18], and high-quality early treatment trials with lopinavir-ritonavir are lacking.

The FLARE trial therefore aimed to deliver robust Phase 2, proof of principle, data on viral load changes using early antiviral treatment. The combination of favipiravir plus lopinavir-ritonavir was studied in a 2×2 factorial design to compare the combination with placebo whilst simultaneously testing each agent in monotherapy to understand their respective contributions. Doses used in current clinical practice and previous trials for other indications were used due to available safety data, and modelling which suggested that we would achieve EC90 for favipiravir based on the available pharmacokinetic data at the time [Supplementary figure 1]. For favipiravir, this is similar to the dose now being employed worldwide for COVID-19.

## Methods

### Study design and participants

FLARE was an early intervention trial testing the effect of oral antiviral therapy on viral load [19]. Participants received favipiravir plus lopinavir-ritonavir, favipiravir plus lopinavir-ritonavir placebo, favipiravir placebo plus lopinavir-ritonavir, or placebos of both drugs. Favipiravir or matched placebo was administered at a dose of 1800 mg twice daily on Day 1, followed by 400 mg four times daily from Day 2 to Day 7. Lopinavir-ritonavir or matched placebo were given at a dose of 400mg/100 mg twice daily on Day 1, followed by 200mg/50mg four times daily from Day 2 to Day 7. Participants were advised to take both Day 1 doses on the first day regardless of time of enrolment, due to the perceived importance of achieving high antiviral levels as early as possible. Those recruited in the afternoon took the first dose immediately and the second dose at least 6 hours later.

Participants aged between 18 and 70 years who had recently (within the last 5 days) developed symptoms of COVID-19, who had tested positive for SARS-CoV-2 by PCR and were within 7 days of symptom onset, or who were asymptomatic but had tested positive by PCR within the previous 48 hours, were eligible for the trial. Participants were ineligible if they had known hypersensitivity to either drug or their ingredients/excipients, had chronic liver or kidney disease, were taking concomitant medicines known to interact with the trial treatments, were being treated as a hospital inpatient for any condition, were pregnant or breastfeeding or were participating in another interventional clinical trial (treatment or vaccination). Before 8 June 2021, participants vaccinated against SARS-CoV-2 were excluded but this was reversed by the Trial Steering Committee due to the large number of vaccinated individuals presenting with infection at that time, and the importance of establishing whether early antiviral treatment is effective in a vaccinated population. Female participants of childbearing potential were required to provide a negative pregnancy test before commencement of trial medication and on Day 14, and to use highly effective contraceptive measures during the trial; male participants with a female partner of childbearing potential were also required to use highly effective contraception.

Participants were informed about the trial via occupational health departments at participating hospital sites and participant identification centres, via poster advertisements, social media or, from 23 June 2021, directly by National Health Service (NHS) Test & Trace following the identification of a positive test. The trial team also directly contacted ambulatory patients who had tested positive at hospital sites and those in the local area from a list provided by NHS Digital.

Participants were recruited at two sites: Royal Free Hospital and University College London Hospital, both in London, UK.

The study was approved by the Wales Research Ethics Committee 3 (Ref: 20/WA/0210) and all participants provided written, informed consent.

### Randomisation and masking

A pre-screening visit (usually by telephone) briefly assessed eligibility and collected the following information: study site, age (≤ 55 vs > 55 years), sex, height and weight (to calculate body mass index (BMI)), symptomatic or asymptomatic, current smoking status (current or non-smoker/ex-smoker), ethnicity, previous COVID-19 specific vaccination (yes/no) and presence/absence of the following comorbidities: diabetes, hypertension, ischaemic or other heart disease or chronic respiratory disease. These variables were used as part of the minimisation strategy to randomise participants into the 4 arms 1:1:1:1 using a centralised concealed online process to assign participants to a medication kit number.

Trial medication kits, prepared by RenaClinical Ltd, were coded to maintain double blinding (investigators and participants). Kits contained favipiravir or colour and size matched placebo 200 mg tablets supplied by Fujifilm Toyama Chemical Co., Ltd and lopinavir-ritonavir 200mg/50 mg tablets (AbbVie Ltd) or colour and size matched placebos (RenaClinical Ltd).

### Procedures

People willing to participate at pre-screening were visited in their home or seen in a designated COVID-19 treatment area at recruitment sites. Following confirmation of eligibility and written informed consent, a nasopharyngeal swab (for participants who were symptomatic but had not tested positive) and baseline blood test was performed along with collection of clinical and demographic information. A pack containing trial medication, kits and instructions for collecting daily saliva samples (Saliva RNA Collection and Preservation devices, Norgen Biotek, Canada), a thermometer and participant diary was provided. The first saliva sample was taken followed by witnessed intake of the first dose of trial medication; participants were advised to take daily saliva samples each morning from Days 2 to 7 before eating, drinking or brushing teeth.

A telephone follow-up was performed on Day 5 and a second visit performed on Day 7 where saliva samples were collected and blood was drawn for safety and favipiravir pharmacokinetics. Stool samples were collected if provided. Follow up telephone calls or visits were made on Day 14; a pregnancy test was performed for women of childbearing potential and blood tests taken if abnormalities had been detected at Day 7. A final telephone call was made on Day 28.

### Outcomes

The primary outcome was viral load measured by quantitative polymerase chain reaction (PCR) performed on saliva samples at Day 5 accounting for the pre-treatment Day 1 viral load. Secondary outcomes were proportion of participants with undetectable viral loads at Day 5, rate of decrease in viral load during the 7-day treatment course, duration of fever, proportion of participants with medication-related toxicity at Days 7 and 14, and proportion of participants admitted to hospital, intensive care or dead due to a COVID-19 related illness.

We planned to assess viral clearance in stool but received insufficient samples for analysis. Further outcomes of whole genome sequencing of SARS-CoV-2 and more extensive pharmacokinetic-pharmacodynamic modelling will be reported separately.

### Laboratory analyses

Full blood count, urea & electrolytes, liver function tests and serum urate were measured in the diagnostic laboratory at Great Ormond Street Hospital (GOSH), London. Saliva viral load was also measured by the GOSH diagnostic laboratory. Samples with a cycle threshold (Ct) value between 40-45 were repeated, and for the purposes of the primary analysis a viral load was calculated from the calibration curve if the repeat value was also <45. However, due to uncertainties in the interpretation of these Ct values and in line with clinical practice, for the secondary analysis of undetectable viral load, Ct values >40 were considered undetectable.

Serum antibody status at Day 1 and Day 7 was measured at the University of Birmingham via enzyme-linked immunosorbent assay, as described previously [20].

Favipiravir drug levels pre and post the second or third dose on Day 7 were measured in plasma by the LSI Medience Corporation in Japan on behalf of Fujifilm Toyama Chemical Co., Ltd. Favipiravir was confirmed to be stable for 24 hours at room temperature and for 6 months once frozen at -20C. The assay lower limit of quantification was 0.1 mg/L.

### Statistical analysis

It was assumed that a clinically significant difference in viral load between antiviral and placebo-treated participants would be 0.5 to 1 log_10_ copies/mL by Day 5. Simulations showed a total of 216 participants would provide 90% power with two-sided alpha of 2.5% to detect a 0.9 log_10_ decrease in viral load of each active treatment on its own compared to placebo. The factorial design allowed an interaction term to be estimated with 80% power, at a nominal two-sided alpha of 5%, to detect a synergistic or antagonistic effect of 1.0 log_10_ copies/mL. To allow for 10% attrition rate a total sample size of 240 (60 participants per arm) was determined.

All statistical analyses were done according to a predefined statistical analysis plan. Analysis of the primary, secondary and safety outcomes was conducted on the intention-to-treat (ITT) population. The ITT population is composed of all randomised participants. For the primary outcome, the ITT analysis was composed of all ITT participants for whom a measure of viral load was available at Day 1 and Day 5. Additionally, the primary outcome was analysed in a modified ITT (mITT) population, which excluded participants who had undetectable viral load both at Day 1 and Day 5.

An analysis of covariance (ANCOVA) model was used to estimate the difference in viral load at 5 days post treatment between the treatment arms. The model included a term for each treatment (favipiravir active/placebo, and lopinavir-ritonavir active/placebo), an interaction term between the two treatments, and baseline viral load. Supportive analyses on the primary outcome included a model adjusting for (i) minimisation factors; (ii) minimisation factors, symptom duration and antibody status (post-hoc adjustment strategy); (iii) potential effect of the delta variant of the SARS-CoV-2 virus, by adding a categorical variable reflecting the period of recruitment: no delta variant (before 24 April 2021), some delta variant (between 24 April 2021 and 12 June 2021) and predominantly delta variant period (post 12 June 2021). A linear mixed model was used to model the viral load trajectories from Day 1 to Day 7 between the four treatment arms. Two adjustment strategies were followed: (i) Day 2 to Day 7 viral loads were modelled as response variable, adjusted for Day 1 viral load; (ii) also adding minimisation factors, symptom duration and antibody status (post-hoc analysis). We used STATA/MP 17·0 for all analyses.

No interim analyses were planned and safety monitoring was undertaken by an Independent Data Monitoring Committee (IDMC). All participants provided written informed consent. The trial registration number was NCT04499677.

## Results

Between 6 October 2020 and 4 November 2021, we screened 1215 and recruited 240 participants (Figure 1). Participant details are provided in Table 1 and minimisation factors in Table 2. Most participants (90%) were below the age of 55 years; 82% were Caucasian and 85% did not have any comorbidities. 51% of those randomised were vaccinated against SARS-CoV-2, and the proportion of vaccinated participants was balanced across the four arms; 63% had detectable SARS-CoV-2 anti-spike antibody at baseline. 66% of the participants started treatment within 5 days of symptom onset. The time between symptom onset and start of treatment was similar between the arms.

**Figure 1.**
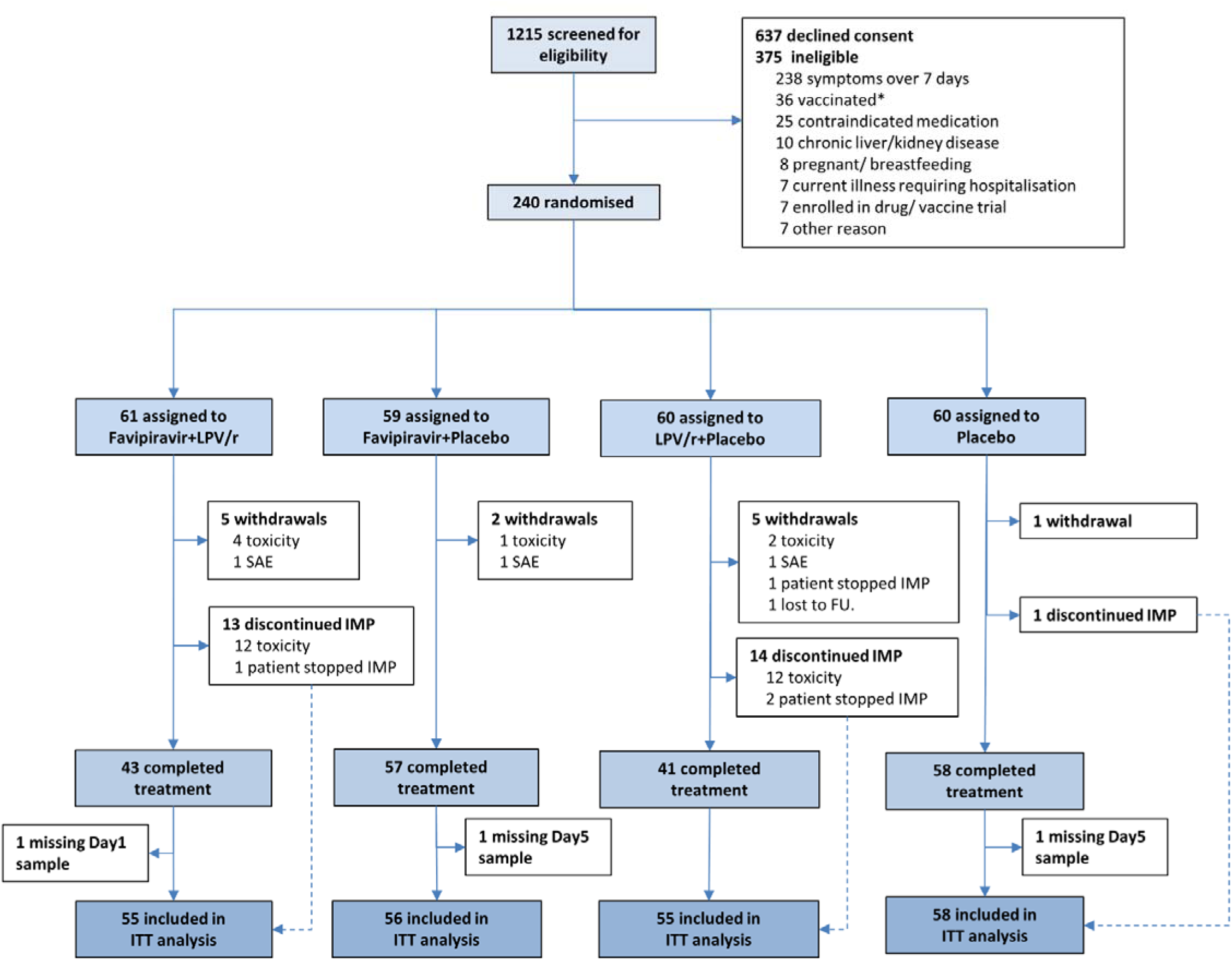
CONSORT diagram for the FLARE trial. * SARS-CoV-2 vaccination was an exclusion in the earlier part of the trial.

**Table 1.** Participant baseline characteristics.

| Characteristics at screening |  | Favipiravir+LPV/r (N=61) | Favipiravir+Placebo (N=59) | LPV/r+Placebo (N=60) | Placebo (N=60) | Total (N=240) |
| --- | --- | --- | --- | --- | --- | --- |
| Age (years) | mean (sd) | 40.3 (13.1) | 40.3 (12.1) | 38.6 (11.5) | 40.6 (12.2) | 40.0 (12.2) |
| Height (cm) | mean (sd) | 172.8 (9.1) | 172.5 (9.6) | 172.1 (9.7) | 171.2 (9.7) | 172.2 (9.5) |
| Weight (kg) | mean (sd) | 76.0 (17.0) | 76.5 (14.1) | 74.8 (16.6) | 75.4 (15.9) | 75.7 (15.9) |
| Pulse Rate (bpm) | mean (sd) | 72.6 (11.4) | 72.6 (11.1) | 76.9 (10.5) | 75.2 (10.9) | 74.3 (11.1) |
| Respiratory Rate (bpm) | mean (sd) | 16.9 (3.5) | 16.5 (2.6) | 16.6 (2.7) | 16.8 (2.8) | 16.7 (2.9) |
| Body Temperature (°C) | mean (sd) | 36.8 (0.7) | 36.7 (0.6) | 36.8 (0.7) | 36.6 (0.6) | 36.7 (0.6) |
| HIV status | N (%) |  |  |  |  |  |
| Positive |  | 1 (1.6) | 0 (0.0) | 0 (0.0) | 0 (0.0) | 1 (0.4) |
| Negative |  | 17 (27.9) | 22 (37.3) | 20 (33.3) | 19 (31.7) | 78 (32.5) |
| Unknown |  | 43 (70.5) | 37 (62.7) | 40 (66.7) | 41 (68.3) | 161 (67.1) |
| Vaccinated | N (%) |  |  |  |  |  |
| Yes |  | 32 (52.5) | 30 (50.8) | 31 (51.7) | 30 (50.0) | 123 (51.2) |
| No |  | 29 (47.5) | 29 (49.2) | 29 (48.3) | 30 (50.0) | 117 (48.8) |
| Type of vaccine | N (%) |  |  |  |  |  |
| Pfizer/BioNTech |  | 14 (23.0) | 13 (22.0) | 19 (31.7) | 8 (13.3) | 54 (22.5) |
| Oxford/AstraZeneca |  | 16 (26.2) | 17 (28.8) | 12 (20.0) | 21 (35.0) | 66 (27.5) |
| Moderna |  | 2 (3.3) | 0 (0.0) | 0 (0.0) | 1 (1.7) | 3 (1.3) |
| Number of doses | N (%) |  |  |  |  |  |
| One |  | 7 (11.5) | 5 (8.5) | 7 (11.7) | 3 (5.0) | 22 (9.2) |
| Two |  | 24 (39.3) | 25 (42.4) | 24 (40.0) | 27 (45.0) | 100 (41.7) |
| Three |  | 1 (1.6) | 0 (0.0) | 0 (0.0) | 0 (0.0) | 1 (0.4) |
| Symptom onset | N (%) |  |  |  |  |  |
| ≤ 5 days |  | 43 (70.5) | 39 (66.1) | 38 (63.3) | 37 (62.7) | 157 (65.7) |
| >5 days |  | 18 (29.5) | 20 (33.9) | 22 (36.7) | 22 (37.3) | 82 (34.3) |
| SARS-CoV-2 antibody status | N (%) |  |  |  |  |  |
| Negative |  | 21 (34.4) | 21 (36.2) | 23 (38.3) | 23 (38.3) | 88 (36.8) |
| Positive |  | 40 (65.6) | 37 (63.8) | 37 (61.7) | 37 (61.7) | 151 (63.2) |

LPV/r: lopinavir-ritonavir, bpm: beats per minute.

**Table 2.** Participant minimisation factors.

| Minimisation factors<br>N (%) | Favipiravir+LPV/r<br>(N=61) | Favipiravir+Placebo<br>(N=59) | LPV/r+Placebo<br>(N=60) | Placebo<br>(N=60) | Total<br>(N=240) |
| --- | --- | --- | --- | --- | --- |
| Site |  |  |  |  |  |
| Royal Free | 56 (91.8) | 55 (93.2) | 55 (91.7) | 55 (91.7) | 221 (92.1) |
| UCLH | 5 (8.2) | 4 (6.8) | 5 (8.3) | 5 (8.3) | 19 (7.9) |
| Age (years) |  |  |  |  |  |
| ≤ 55 | 53 (86.9) | 52 (88.1) | 55 (91.7) | 55 (91.7) | 215 (89.6) |
| > 55 | 8 (13.1) | 7 (11.9) | 5 (8.3) | 5 (8.3) | 25 (10.4) |
| Gender |  |  |  |  |  |
| Male | 31 (50.8) | 32 (54.2) | 29 (48.3) | 31 (51.7) | 123 (51.2) |
| Female | 30 (49.2) | 27 (45.8) | 31 (51.7) | 29 (48.3) | 117 (48.8) |
| Ethnicity |  |  |  |  |  |
| Caucasian | 50 (82.0) | 49 (83.1) | 49 (81.7) | 49 (81.7) | 197 (82.1) |
| Other | 11 (18.0) | 10 (16.9) | 11 (18.3) | 11 (18.3) | 43 (17.9) |
| BMI (kg/m<sup>2</sup>) |  |  |  |  |  |
| <30 | 51 (83.6) | 49 (83.1) | 50 (83.3) | 50 (83.3) | 200 (83.3) |
| ≥30 | 10 (16.4) | 10 (16.9) | 10 (16.7) | 10 (16.7) | 40 (16.7) |
| Symptomatic disease |  |  |  |  |  |
| Yes | 61 (100.0) | 59 (100.0) | 60 (100.0) | 59 (98.3) | 239 (99.6) |
| No | 0 (0.0) | 0 (0.0) | 0 (0.0) | 1 (1.7) | 1 (0.4) |
| Current smoker |  |  |  |  |  |
| Yes | 6 (9.8) | 7 (11.9) | 7 (11.7) | 7 (11.7) | 27 (11.3) |
| No | 55 (90.2) | 52 (88.1) | 53 (88.3) | 53 (88.3) | 213 (88.8) |
| Vaccinated |  |  |  |  |  |
| Yes | 32 (52.5) | 30 (50.8) | 31 (51.7) | 30 (50.0) | 123 (51.2) |
| No | 29 (47.5) | 29 (49.2) | 29 (48.3) | 30 (50.0) | 117 (48.8) |
| Comorbidity |  |  |  |  |  |
| Present | 11 (18.0) | 9 (15.3) | 8 (13.3) | 8 (13.3) | 36 (15.0) |
| Absent | 50 (82.0) | 50 (84.7) | 52 (86.7) | 52 (86.7) | 204 (85.0) |

As detailed in Figure 1, 13 participants withdrew from the trial and a further 28 discontinued medication but provided samples for analysis. Predominantly this was due to toxicity which occurred disproportionately in arms including lopinavir-ritonavir (see Safety below). Overall 224 participants (93.3%) were included in the ITT analysis and 208 participants (86.7%) in the mITT analysis of the primary outcome.

The primary outcome was SARS-CoV-2 viral load at Day 5 of therapy accounting for baseline viral load. Figure 2 and Table 3 present summary data for the entire ITT and mITT cohorts, while Supplementary Figure 2 displays results at participant level. In the primary analysis, there was no significant effect of any treatment arm on viral load: additional reduction in viral load versus placebo for favipiravir monotherapy 0.57 log_10_ copies/mL (95% confidence interval (CI) -1.21 to 0.07, p=0.08), for lopinavir-ritonavir monotherapy 0.18 log_10_ copies/mL (95% CI -0.82 to 0.46, p=0.58). There was no significant interaction between favipiravir and lopinavir-ritonavir but the coefficient was numerically in the direction of antagonism (interaction coefficient: 0.59 log_10_ copies/mL, 95% CI - 0.32 to 1.50, p=0.20).

**Figure 2.**
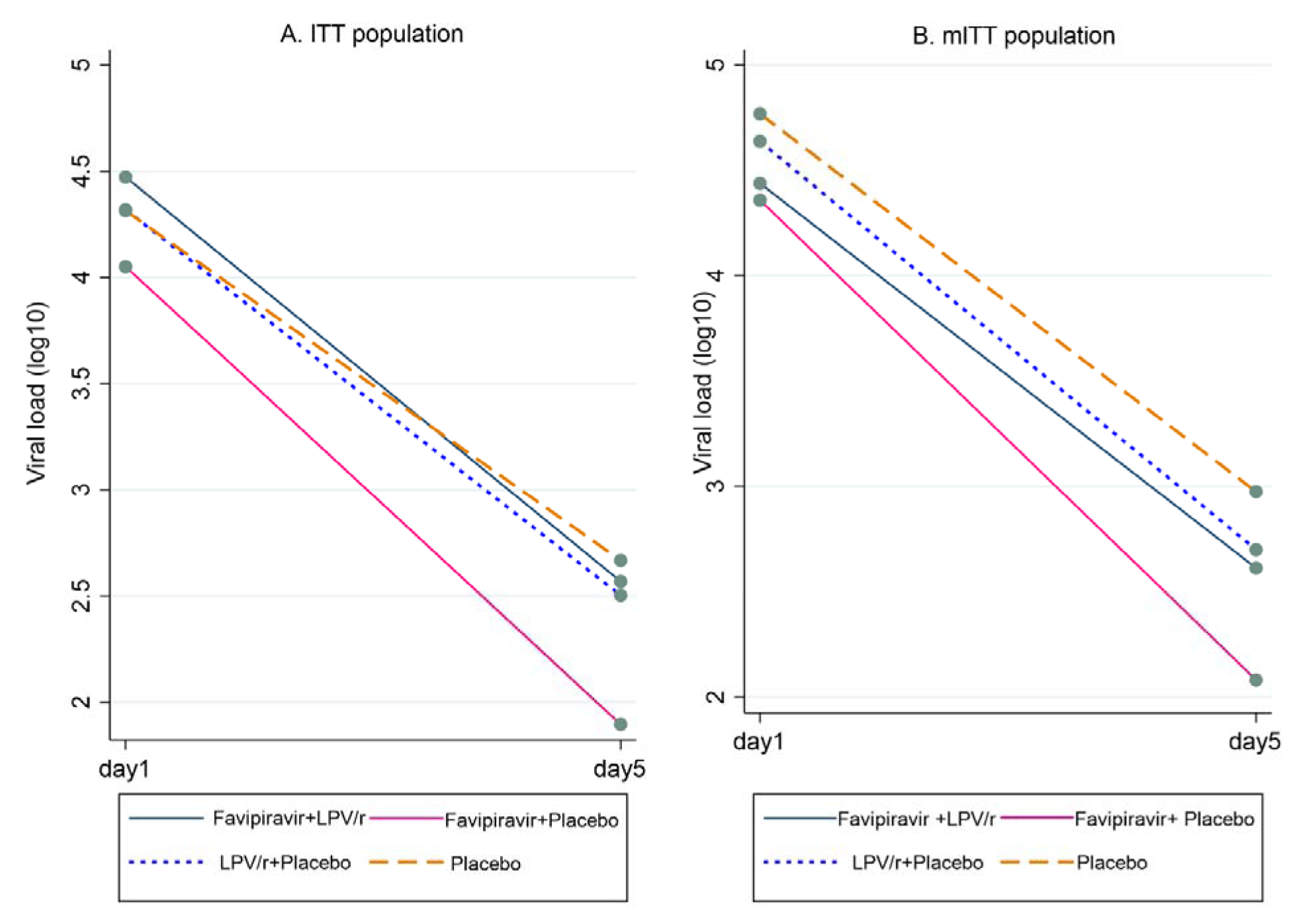
Mean log_10_ SARS-CoV-2 viral load at baseline (Day 1) and Day 5 per treatment arm in (A) the full intention to treat (ITT) population and (B) the modified intention to treat (mITT) population, excluding participants with negative viral load at baseline and Day 5.

**Table 3.** Primary outcome analysis: SARS-CoV-2 viral load at Day 5 adjusted for baseline viral load.

|  | N | Favipiravir+Placebo<br>(Main effect) |  | LPV/r+Placebo<br>(Main effect) |  | Interaction<br>Favipiravir+LPV/r |  |
| --- | --- | --- | --- | --- | --- | --- | --- |
|  |  | Coefficient<br>(95% CI) | p-value | Coefficient<br>(95% CI) | p-value | Coefficient<br>(95% CI) | p-value |
| Primary outcome |  |  |  |  |  |  |  |
| ITT population | 224 | -0.57<br>(-1.21, 0.07) | 0.08 | -0.18<br>(-0.82, 0.46) | 0.58 | 0.59<br>(-0.32, 1.50) | 0.20 |
| Modified ITT population | 208 | -0.59<br>(-1.29, 0.11) | 0.10 | -0.18<br>(-0.87, 0.51) | 0.61 | 0.65<br>(-0.33, 1.63) | 0.19 |
| Adjusted analyses of primary outcome |  |  |  |  |  |  |  |
| Adjusted for minimisation factors | 224 | -0.57<br>(-1.16, 0.02) | 0.06 | -0.14<br>(-0.73, 0.45) | 0.65 | 0.62<br>(-0.22, 1.46) | 0.15 |
| Adjusted for minimisation factors,<br>symptom duration, antibody status | 222 | -0.65 (-1.23, -<br>0.07) | 0.03 | -0.09 (-0.66, 0.49) | 0.76 | 0.66 (-0.16, 1.48) | 0.11 |
| Mixed model analysis - At day 5 |  |  |  |  |  |  |  |
| ITT population | 235 | -0.57 (-1.14,<br>0.01) | 0.05 | -0.24 (-0.81, 0.34) | 0.43 | 0.65 (-0.17, 1.47) | 0.12 |
| Adjusted for minimisation factors,<br>symptom duration, antibody status | 233 | -0.63 (-1.17, -<br>0.08) | 0.02 | -0.15 (-0.69, 0.40) | 0.60 | 0.65 (-0.11, 1.42) | 0.10 |

For favipiravir monotherapy, we observed similar effect sizes after adjustment for minimisation factors or for a potential effect of the delta variant of SARS-CoV-2 (p=0.06). However, adjusting for the minimisation factors as well as symptom duration and antibody status, a stronger effect was noted (−0.65 log_10_ copies/mL [95% CI -1.23 to -0.07], p=0.03). Following the same adjustment strategy and conditioning on baseline viral load, the mixed model analysis indicated a similar effect of favipiravir monotherapy that reached our pre-defined threshold for significance (−0.63 log_10_ copies/mL [95% CI -1.17 to -0.08], p=0.02; Table 3).

The proportion of participants with undetectable viral load at Day 5 was somewhat higher in the favipiravir monotherapy arm (odds ratio of being undetectable 2.47 [95% CI 1.08 to 5.65, p=0.03]) but there was no effect of other treatment arms (Table 4).

**Table 4.** Odds ratios of achieving undetectable viral load (Ct ≥40) by Day 5.

|  | Sample size* | Placebo | Favipiravir+ Placebo<br>(Main effect) |  |  | LPV/r+Placebo<br>(Main effect) |  |  | Interaction<br>Favipiravir+LPV/r |  |  |
| --- | --- | --- | --- | --- | --- | --- | --- | --- | --- | --- | --- |
|  |  | N (%) | N (%) | OR<br>(95% CI) | p-value | N (%) | OR<br>(95% CI) | p-value | N (%) | OR<br>(95% CI) | p-value |
| Undetectable viral load | 203 | 14<br>(26.9) | 25<br>(46.3) | 2.47<br>(1.08, 5.65) | 0.03 | 17<br>(30.4) | 1.29<br>(0.55, 3.00) | 0.56 | 20<br>(35.7) | 0.52<br>(0.16, 1.66) | 0.27 |

\* Patients included in this analysis had a detectable viral load at baseline and saliva sample available at Day 5.
LPV/r: lopinavir-ritonavir

In post-hoc supportive analyses, we observed a significant interaction (p=0.03) between treatment with favipiravir and baseline viral load levels (above or below the median level of 4.56 log_10_ copies/mL). In the low viral load group, there was no difference in Day 5 viral load between the treatment arms. However, in the high viral load group, favipiravir monotherapy was associated with a reduced viral load compared to placebo at Day 5 (difference 1.30 log_10_ copies/mL [95% CI 0.30 to 2.29]; Figure 3 and Table 5).

**Figure 3.**
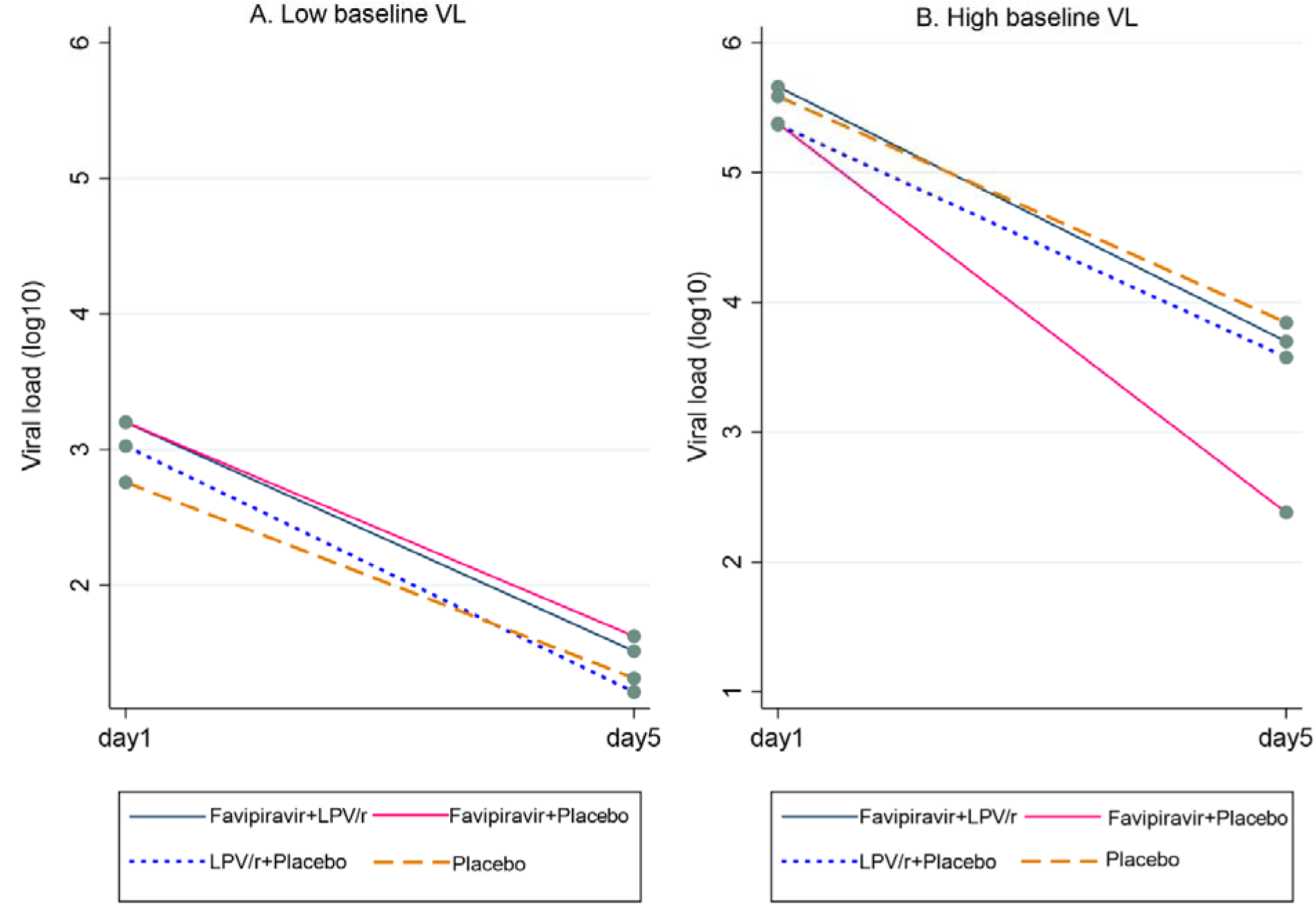
Mean log_10_ SARS-CoV-2 viral load at baseline (Day 1) and Day 5 per treatment arm in (A) participants with baseline viral load below or equal to the median level for the entire cohort and (B) participants with baseline viral load above the median level for the entire cohort.

**Table 5.** Subgroup analyses for primary outcome according to vaccination status, duration of symptoms, baseline antibody status and baseline viral load.

|  | N | Placebo | Favipiravir+Placebo<br>(Main effect) |  |  | LPV/r+Placebo<br>(Main effect) |  |  | Interaction<br>Favipiravir+LPV/r |  |  |
| --- | --- | --- | --- | --- | --- | --- | --- | --- | --- | --- | --- |
|  |  | N | N | Coefficient<br>(95% CI) | Interaction<br>p-value | N | Coefficient<br>(95% CI) | Interaction<br>p-value | N | Coefficient<br>(95% CI) | Interaction<br>p-value |
| Vaccinated |  |  |  |  |  |  |  |  |  |  |  |
| Yes | 117 | 29 | 28 | -0.71 (-1.66, 0.24) | 0.67 | 29 | 0.15 (-0.79, 1.09) | 0.32 | 31 | 0.90 (-0.43, 2.23) | 0.57 |
| No | 107 | 29 | 28 | -0.41 (-1.09, 0.27) |  | 26 | -0.45 (-1.14, 0.24) |  | 24 | 0.36 (-0.64, 1.35) |  |
| Days from symptom onset |  |  |  |  |  |  |  |  |  |  |  |
| ≤ 5 days | 148 | 35 | 38 | -0.37 (-1.17, 0.44) | 0.55 | 35 | 0.02 (-0.79, 0.84) | 0.50 | 40 | 0.48 (-0.65, 1.61) | 0.93 |
| >5 days | 75 | 22 | 18 | -0.80 (-1.86, 0.26) |  | 20 | -0.43 (-1.46, 0.60) |  | 15 | 0.42 (-1.13, 1.97) |  |
| Baseline antibody status |  |  |  |  |  |  |  |  |  |  |  |
| Negative | 80 | 23 | 20 | -0.06 (-0.75, 0.63) | 0.27 | 20 | -0.13 (-0.81, 0.55) | 0.98 | 17 | -0.09 (-1.10, 0.91) | 0.24 |
| Positive | 143 | 35 | 35 | -0.86 (-1.72, -0.01) |  | 35 | -0.14 (-1.0, 0.72) |  | 38 | 1.08 (-0.11, 2.28) |  |
| Baseline viral load |  |  |  |  |  |  |  |  |  |  |  |
| ≤ Median viral load | 117 | 27 | 36 | 0.12 (-0.72, 0.96) | 0.03 | 35 | -0.20 (-1.11, 0.70) | 0.94 | 29 | 0.09 (-1.13, 1.31) | 0.17 |
| >Median viral load | 107 | 31 | 20 | -1.30 (-2.29, -0.30) |  | 30 | -0.13 (-1.01, 0.76) |  | 26 | 1.28 (-0.09, 2.65) |  |

LPV/r: lopinavir-ritonavir

We also analysed results according to pre-specified subgroups (vaccination status, antibody status and duration of symptoms before commencing treatment (≤5 days versus >5 days)) but did not observe any differences between treatments across subgroups (Table 5).

Finally, we plotted average viral load in the ITT population (also dividing into high and low baseline viral load groups) and proportion with undetectable viral load per day of treatment (Supplementary Figures 3 and 4). Broadly, similar patterns were observed throughout the treatment course. Of note, we observed steeper decline of viral load in vaccinated or antibody-positive participants, with somewhat lower baseline viral loads in the latter, regardless of treatment arm (Supplementary Figure 5).

A total of 518 adverse events were reported in 191 (80%) participants, of which 295 (57%) events were considered related to the treatment. The proportion of participants with treatment-related events was greater in those receiving lopinavir-ritonavir monotherapy (93%) and favipiravir plus lopinavir-ritonavir (88%) compared to those receiving favipiravir monotherapy (46%) and placebo (35%). The odds of experiencing a related event were significantly higher in the lopinavir-ritonavir arm compared to placebo (OR 16.0 [95% CI 4.27 to 60.0], p<0.0001). Specifically, the number of events of diarrhoea and nausea was higher in participants treated with lopinavir-ritonavir and combination therapy. As detailed above, more participants in arms containing lopinavir-ritonavir discontinued treatment. Adverse events are summarised in Supplementary Table 1.

We also measured liver function tests at Day 1 and Day 7 (Supplementary Table 2 and Supplementary Figure 6). Median levels for all parameters were within the normal range at both time points with minimal change during treatment. No clinically significant hepatitis or other hepatotoxicity was observed, but a minority of participants had a mild transaminitis before or during treatment. Participants with abnormal tests had repeat samples on Day 14 (Supplementary Figure 5).

As expected, serum uric acid levels significantly increased in the arms containing favipiravir (odds ratio for elevated uric acid level in favipiravir monotherapy arm 18.8 [95% CI 4.2 to 84.8], p<0.0001) after seven days of treatment. However, the high levels were not sustained at Day 14.

There were three serious adverse events during the trial, all were hospitalisation due to progression of COVID-19. One event was seen in each of the lopinavir-ritonavir monotherapy, favipiravir monotherapy and combination treatment arms. One participant (in the favipiravir monotherapy arm) was admitted to intensive care. There were no deaths in the study.

All participants still taking trial medication and who were seen on Day 7 had blood samples taken pre-dose and 30-60 minutes post-dose for measurement of favipiravir drug levels. Assays were run on samples from 31 participants in the favipiravir monotherapy arm and 28 participants in the combination arm. As shown in Figure 4, favipiravir levels at both trough and peak were significantly lower in the combination treatment arm than in the favipiravir monotherapy arm. Of note, only a minority of participants achieved levels close to the EC90. Supplementary Table 3 summarises demographic data on this cohort of participants, which did not differ between the arms or from the overall characteristics of the participants randomised to these arms.

**Figure 4.**
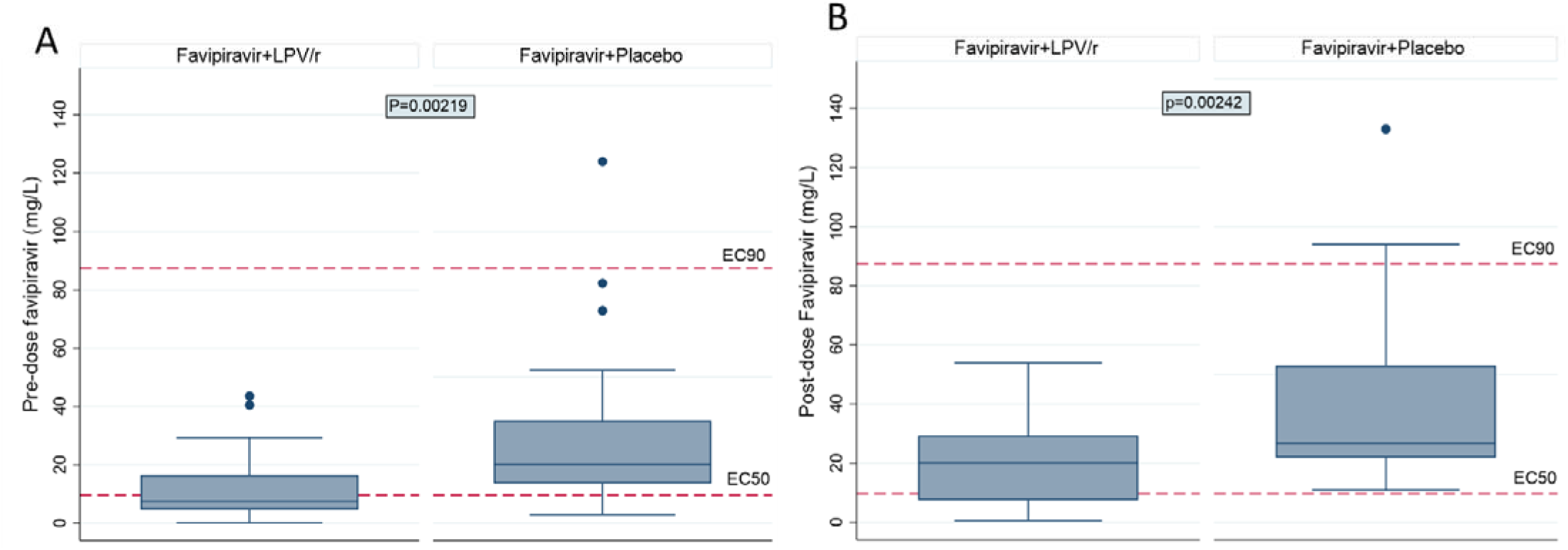
Plasma favipiravir concentration in the combination favipiravir + lopinavir-ritonavir (LPV/r) arm and the favipiravir + placebo arm on Day 7 (A) pre-dose (trough) and (B) 30-60 minutes post-dose (peak).

There was no difference in duration of fever between the arms, which was only observed in a minority of participants. There were also no differences between the arms in the proportion of participants with positive anti-spike antibody by Day 7, quantitative antibody levels or the magnitude of change from Day 1.

## Discussion

When the FLARE trial was designed in March 2020, we identified the imperative to generate high-quality Phase 2 proof of principle trial evidence on repurposed antivirals for early treatment of COVID-19, and this question remains important. The trial opened for recruitment in September 2020, but proceeded predominantly at a single site as we did not receive research prioritisation in the UK via Urgent Public Health (UPH) status.

Based on *in vitro* data and early clinical reports, favipiravir was chosen as the most promising orally available agent. Due to uncertainty whether favipiravir would be effective as monotherapy, the addition of lopinavir-ritonavir was proposed as an inexpensive, readily available protease inhibitor with evidence of some clinical effect against previous coronaviruses and modest *in vitro* anti-SARS-CoV-2 activity. The major finding of FLARE is that, at the doses used, there is no clear evidence that either favipiravir monotherapy or favipiravir plus lopinavir-ritonavir produce clinically worthwhile reductions in viral load in early treatment. FLARE provides insufficient evidence to take favipiravir monotherapy or favipiravir plus lopinavir-ritonavir into Phase 3, and instead predicts that none of the intervention arms would provide important clinical benefit at the current dose. However, further study of favipiravir may be warranted. In particular, dose escalation studies might potentially identify more efficacious doses against SARS-CoV-2.

We found a numerically greater but non-significant reduction in viral load associated with favipiravir monotherapy in the primary analysis, while a post-hoc fully adjusted mixed model, similar to that used to report the effect of other antivirals [21], was modestly statistically significant at our pre-specified threshold (Table 3, Figure 2). We also observed an increase in the proportion of patients with undetectable viral load compared to placebo, lopinavir-ritonavir or combination therapy (Table 4, Supplementary Figure 4). The effect was seen especially in those with higher baseline viral load, likely due to viral replication having slowed substantially in those with low viral load, limiting the potential for antivirals to inhibit replication [17, 18]. However, this may point towards efficacy in a group with the most potential to benefit.

Favipiravir is a ribosomal-dependent RNA polymerase (RdRp) inhibitor with a similar mode of action to molnupiravir. The magnitude of difference in viral load at Day 5 with favipiravir versus placebo in our trial (0.57 to 0.65 log_10_ copies/mL, depending on analysis used) was similar to that seen with molnupiravir (0.55 log_10_ copies/mL) at the highest dose tested in trials (800mg twice daily) [21]: this agent has been reported to be clinically effective for early COVID-19. However, it remains to be seen whether molnupiravir monotherapy will retain clinical benefits in routine clinical practice. Favipiravir as monotherapy was well tolerated with relatively few adverse effects; in particular, we did not observe significant hepatotoxicity indicating that it may be well tolerated at higher doses. Indeed, a loading dose of 6000 mg (2400 mg given twice 8 hours apart followed by 1200 mg) on Day 1, followed by 1200 mg twice daily thereafter was well tolerated when used in Ebola [22]. High levels of uric acid were seen, which is a well-recognised side effect of favipiravir, but without obvious clinical consequence.

We chose the favipiravir dose used in influenza trials of 3600 mg on Day 1 followed by 1600 mg daily thereafter because simulations using pharmacokinetic data provided by Fujifilm Toyama Chemical Co., Ltd suggested we should expect to achieve 90% viral replication inhibition (along with a slight advantage in higher pre-dose trough levels if the maintenance dose was split 4 times per day rather than twice per day [Supplementary Figure 1]). However, upon measuring favipiravir pharmacokinetics on Day 7, we found levels around one third of our pre-trial predictions and, perhaps more unexpectedly, significantly lower levels of favipiravir in the combination arm despite measurement being limited to those still taking IMP at this time point (Figure 4).

Our dosing simulations assumed linear pharmacokinetics and although there was a prior report of time-dependent reductions in levels seen in Ebola [22], it was not clear that this would be the case with our dose regimen. However, pharmacokinetic data published after the start of FLARE indicate that favipiravir is likely to display time-dependent nonlinear pharmacokinetics at the doses used here [23], albeit intracellular concentrations with this dose regimen have been proposed to reach antiviral levels [24]. However, this time-dependent nonlinearity does not account for the lower levels seen in the combination compared with monotherapy arm. Whilst a cytochrome P450 mediated drug-drug interaction is not expected between favipiravir and lopinavir-ritonavir, possible explanations include lower favipiravir absorption associated with the gastrointestinal effects of lopinavir-ritonavir, or more unreported missed doses in the combination arm.

It remains possible that a concentration-dependent antiviral effect may nevertheless occur with the lower concentrations seen in FLARE, especially via mutagenesis. Viral sequencing work is ongoing to explore this possibility and a population pharmacokinetic-pharmacodynamic model is planned to investigate whether there is a concentration-response relationship with either viral load or mutagenesis. This model should identify the rationale for and doses to use in a future Phase 2 trial.

By including a placebo-controlled lopinavir-ritonavir monotherapy arm, FLARE has demonstrated that this agent has no potential to reduce viral load and is poorly tolerated particularly when treatment is first initiated. As such, FLARE provides a strong rationale not to take this drug into Phase 3. We were able to reach this conclusion by exposing only 60 outpatients to lopinavir-ritonavir monotherapy. A similar design could have quickly ruled out other repurposed agents such as hydroxychloroquine.

An expected but problematic issue encountered with lopinavir-ritonavir was the frequency of side effects, especially gastrointestinal, leading to frequent discontinuation of treatment. We also encountered numerous potential drug-drug interactions, including with commonly prescribed medications such as budesonide and simvastatin, requiring exclusion of potential participants or modification/suspension of concomitant medications. These are important issues to consider with other ritonavir-boosted protease inhibitors (e.g. nirmatrelvir).

As a result of the prolonged recruitment period for FLARE which coincided with the successful UK vaccine roll-out, the Trial Steering Committee decided to include participants who had received a vaccine. Regardless of treatment arm, rate of viral load decay tended to be higher in participants who were vaccinated or antibody-positive at baseline.

Favipiravir is in routine usage for COVID-19 in many countries, but existing trial data are mixed. Some small, open-label studies have indicated benefits in terms of clinical outcomes [25–28] or viral shedding [13, 26]. However, other studies have indicated no clinically important benefit [29, 30], including when given in early disease [31]. These studies were open label with heterogenous populations often including hospitalised patients, where antiviral treatment is expected to be less effective. Holubar *et al* performed a double-blind randomized trial of favipiravir in asymptomatic or mildly symptomatic adults within 72 hours of a positive SARS-CoV2 RT-PCR (median 5 days of symptoms) [14]. Among 116 patients, there was no difference in time to viral shedding cessation or symptom resolution. However, baseline Ct value (inversely related to viral load) tended to be lower while the change in Ct value between Days 1 to 7 tended to be greater in the favipiravir-treated arm.

Our study has some limitations. The recruited cohort was relatively young and healthy with lower viral loads than many reported elsewhere in the literature. We were unable to perform viral culture or infectivity assays which may have provided useful additional information. For logistical reasons, we were unable to obtain samples for pharmacokinetics on every participant in the study.

In conclusion, our results do not support routine usage or Phase 3 trials of favipiravir or lopinavir-ritonavir at the doses investigated. However, the results may indicate an effect of favipiravir when used for early treatment of COVID-19, especially in those with high baseline viral load, but further investigation is needed regarding dosing schedule or additive medication. Another relatively small study would be sufficient to establish this. We have conclusively demonstrated the ineffectiveness of lopinavir-ritonavir even in early disease and have identified a new drug interaction between favipiravir and lopinavir-ritonavir with the latter apparently lowering plasma levels of the former. These results have important implications for the global efforts against COVID-19.

## Data Availability

All data produced in the present study are available upon reasonable request to the authors

## Contributions

Conceptualisation: DML, H-MD, JB, NF, JFS. Formal analysis: KC, AAA, H-MD, NF, JFS. Funding acquisition: DML, JB, NF, JFS. Investigation: DML, LKB, SD, PY, DS, AL, AR, NL, AC. Methodology: DML, H-MD, JB, NF, JFS. Project administration: L-KB, FI, AN. Writing – original draft: DML, JFS. Writing – review & editing: all authors.

## Acknowledgements

The authors would like to acknowledge: the Agile Lighthouse team within UKHSA for assistance with recruitment and Fujifilm Toyama Chemical Co. who provided favipiravir and favipiravir placebo free of charge. We also acknowledge Professor Chris Frost as a non-independent member of the Trial Steering Committee and Dr Mak Wenyao for intellectual contribution. The study was funded by LifeArc (COVID0005). JFS was funded by a UK Medical Research Council (MRC) fellowship (MR/M008665/) and AAA was supported by a MRC project grant (MR/W015560/1).

---

**Supplementary Table 1.** Summary of adverse events.

|  | **Favipiravir+LPV/r**<br>**(N=61)** | **Favipiravir+Placebo**<br>**(N=59)** | **LPV/r+Placebo**<br>**(N=60)** | **Placebo**<br>**(N=60)** | **Total**<br>**(N=240)** |
| --- | --- | --- | --- | --- | --- |
| **Number of Patients reporting at least 1 AE; N (%)** | 55 (90.1) | 38 (64.4) | 59 (98.3) | 39 (65.0) | 191 (80.0) |
| **Patients with at least one related event** | 53 (87.9) | 27 (45.8) | 56 (93.3) | 21 (35.0) | 157 (65.4) |
| **Number of AEs** | 159 | 92 | 175 | 92 | 518 |
| **Related events** | 108 (67.9) | 44 (47.3) | 116 (65.9) | 27 (29.3) | 295 (56.7) |
| **AE Event [# events]** |  |  |  |  |  |
| Diarrhoea | 41 | 8 | 47 | 10 | 106 |
| Nausea | 16 | 13 | 28 | 6 | 63 |
| Dyspnea | 5 | 6 | 7 | 6 | 24 |
| Headache | 6 | 7 | 4 | 6 | 23 |
| Anosmia | 5 | 3 | 9 | 5 | 22 |
| Fatigue | 4 | 4 | 7 | 7 | 22 |
| Vomiting | 8 | 1 | 6 | 2 | 17 |
| Cough | 2 | 5 | 4 | 5 | 16 |
| Dysgeusia | 3 | 4 | 6 | 3 | 16 |
| Abdominal pain | 2 | 3 | 2 | 5 | 12 |
| Anorexia | 2 | 1 | 7 | 2 | 12 |
| Dizziness | 4 | 1 | 6 | 0 | 11 |
| Alanine aminotransferase increased | 6 | 1 | 1 | 1 | 9 |
| Myalgia | 3 | 2 | 1 | 3 | 9 |
| Rash maculo-papular | 2 | 4 | 1 | 2 | 9 |
| Aspartate aminotransferase increased | 4 | 0 | 1 | 2 | 7 |
| Nasal congestion | 1 | 3 | 1 | 2 | 7 |
| Non-cardiac chest pain | 4 | 0 | 1 | 2 | 7 |
| Hyperuricemia | 0 | 2 | 0 | 0 | 2 |

**Supplementary Table 2.** Serum liver function tests and uric acid at Day 1 and Day 7.

|  | Favipiravir+LPV/r<br>(N=61) |  | Favipiravir+Placebo<br>(N=59) |  | LPV/r+Placebo<br>(N=60) |  | Placebo<br>(N=60) |  | Total<br>(N=240) |  |
| --- | --- | --- | --- | --- | --- | --- | --- | --- | --- | --- |
|  | median<br>(IQR) | Outside<br>normal<br>range<br>N (%) | median<br>(IQR) | Outside<br>normal<br>range<br>N (%) | median<br>(IQR) | Outside<br>normal<br>range<br>N (%) | median<br>(IQR) | Outside<br>normal<br>range<br>N (%) | median<br>(IQR) | Outside<br>normal<br>range<br>N (%) |
| **ALT (IU/L)** |  |  |  |  |  |  |  |  |  |  |
| Day 1 | 27.0 (19.0-41.0) | 18 (30.0) | 28.0 (20.0-45.0) | 15 (25.9) | 24.0 (15.0-36.5) | 10 (16.9) | 24.5 (15.0-38.5) | 15 (25.0) | 26.0 (16.0-41.0) | 58 (24.5) |
| Day 7 | 27.0 (18.0-37.0) | 15 (25.0) | 35.5 (24.5-50.5) | 22 (37.9) | 21.0 (15.0-28.0) | 4 (6.8) | 22.5 (17.5-36.0) | 13 (21.7) | 26.0 (18.0-38.5) | 54 (22.8) |
| Change | -2.0 (-12.0- 6.0) |  | 1.0 (-3.0-12.0) |  | -2.0 (-8.0- 1.0) |  | 0.0 (-4.5- 5.0) |  | -1.0 (-7.0- 5.0) |  |
| **AST (IU/L)** |  |  |  |  |  |  |  |  |  |  |
| Day 1 | 32.0 (28.0-41.0) | 18 (30.0) | 34.0 (28.0-43.0) | 19 (32.8) | 31.0 (27.0-36.5) | 12 (20.3) | 30.5 (26.0-36.5) | 16 (26.7) | 32.0 (27.0-38.0) | 65 (27.4) |
| Day 7 | 28.0 (25.0-34.0) | 13 (21.7) | 32.5 (28.0-39.5) | 15 (25.9) | 28.0 (25.0-31.0) | 3 (5.1) | 28.0 (24.0-32.5) | 9 (15.0) | 29.0 (25.0-34.0) | 40 (16.9) |
| Change | -3.0 (-8.0- 1.0) |  | -2.0 (-6.0- 3.0) |  | -2.0 (-6.0- 0.0) |  | -1.5 (-5.0- 1.5) |  | -2.0 (-7.0- 2.0) |  |
| **ALP (IU/L)** |  |  |  |  |  |  |  |  |  |  |
| Day 1 | 60.0 (53.0-74.0) | 1 (1.7) | 60.5 (54.0-70.0) | 0 (0.0) | 57.5 (49.0-72.5) | 0 (0.0) | 60.5 (52.0-72.5) | 1 (1.7) | 60.0 (52.0-72.0) | 2 (0.8) |
| Day 7 | 67.0 (57.0-83.0) | 2 (3.3) | 66.0 (58.5-77.0) | 1 (1.7) | 60.0 (51.0-76.0) | 1 (1.7) | 66.0 (54.5-77.0) | 1 (1.7) | 65.0 (55.0-78.0) | 5 (2.1) |
| Change | 4.0 (1.0-12.0) |  | 6.0 (2.0-13.0) |  | 2.0 (-2.0- 6.0) |  | 1.5 (-1.0- 8.0) |  | 4.0 (0.0-10.0) |  |
| **Bilirubin (μmol/L)** |  |  |  |  |  |  |  |  |  |  |
| Day 1 | 5.0 (4.0- 8.0) | 1 (1.7) | 6.0 (4.0- 8.0) | 1 (1.7) | 6.0 (3.0- 7.0) | 0 (0.0) | 7.0 (4.0- 9.0) | 0 (0.0) | 6.0 (4.0- 8.0) | 2 (0.8) |
| Day 7 | 10.0 (6.0-14.0) | 2 (3.3) | 6.0 (4.0- 9.0) | 1 (1.7) | 8.5 (6.0-13.0) | 4 (6.8) | 7.0 (5.0-10.0) | 1 (1.7) | 8.0 (5.0-12.0) | 8 (3.4) |
| Change | 4.0 (1.0- 8.0) |  | 1.0 (-1.0- 3.0) |  | 4.5 (0.0- 8.5) |  | 2.0 (-2.0- 3.0) |  | 2.0 (0.0- 5.0) |  |
| **Uric acid (μmol/L)** |  |  |  |  |  |  |  |  |  |  |
| Day 1 | 275.0 (209.0-336.0) | 7 (11.5) | 253.0 (216.0-315.0) | 5 (8.5) | 256.5 (196.0-297.5) | 6 (10.0) | 285.0 (210.5-327.0) | 5 (8.3) | 265.0 (209.0-320.0) | 23 (9.6) |
| Day 7 | 369.5 (299.0-441.0) | 18 (29.5) | 422.5 (349.5-498.0) | 22 (37.3) | 258.5 (203.5-316.0) | 5 (8.3) | 275.5 (238.5-346.0) | 2 (3.3) | 329.0 (251.0-401.0) | 47 (19.6) |
| Day 14 | 306.0 (265.0 - 400.0) | 2 (8.0) | 329.0 (287.0 - 354.0) | 1 (3.7) | 288.5 (247.5 - 331.5) | 1 (5.0) | 310.0 (237.0 - 353.0) | 2 (9.1) | 303.5 (250.0 - 253.0) | 6 (6.4) |
| Change | -2.0 (-12.0-6.0) |  | 1.0 (-3.0-12.0) |  | -2.0 (-8.0- 1.0) |  | 0.0 (-4.5- 5.0) |  | -1.0 (-7.0- 5.0) |  |

LPV-r: lopinavir/ritonavir, ALT: alanine aminotransferase, AST: aspartate aminotransferase, ALP: alkaline phosphatase, IU: international units, $\mu$ mol: micromoles

**Supplementary Table 3.** Baseline characteristics of the cohort with pharmacokinetic measurements.

| Characteristics of Pk group at screening |  | Favipiravir+LPV/r (N=28) | Favipiravir+Placebo (N=31) |
| --- | --- | --- | --- |
| Age (years) | mean (sd) | 39.4 (13.4) | 40.9 (11.7) |
| Gender | N (%) |  |  |
| Male |  | 16 (57.1) | 17 (54.8) |
| Female |  | 12 (42.9) | 14 (45.2) |
| Ethnicity | N (%) |  |  |
| Caucasian |  | 23 (82.1) | 27 (87.1) |
| Other |  | 5 (17.9) | 4 (12.9) |
| BMI (kg/m<sup>2</sup>) | N (%) |  |  |
| <30 |  | 23 (82.1) | 49 (83.1) |
| ≥30 |  | 5 (17.9) | 5 (16.1) |

## Supplementary Appendix – FLARE Investigators

**Trial Steering Committee:**

Kristina Nadrah (Chair), Robert C Read, Elizabeth Allen, Mahdia Sait

**Independent Data Monitoring Committee:**

Stephen Owens (Chair), David Chadwick, April Slee, Andrew Ustianowski

**UCL Comprehensive Clinical Trials Unit:**

Krishneya Anojan, Gemma Jones, Nazma Begum-Ali, Natasha Majid

**Royal Free Hospital Clinical Trials team:**

Rachel Ochiel, Debbie Falconer, Stella O’Connor, Karl Salazar, Tung Le, Francesca Gowing, Ivy Wanjiku Dakouri, Tanaka Ngcozana, Sandra Lopez Garces, Karima Oduka, Daniel Jones, Eva Torok-Pollok

**University College London Hospital Clinical Trials team:**

Michelle Berkeley, Esther King, Kimberlee Gunn

**Great Ormond Street Hospital laboratories:**

Francis Yongblah, Mabel Csatari, Kimberly Gilmour

**Royal Free Hospital and UCL (Royal Free campus) laboratories:**

Naseem Ahmed, Janki Kavi, Nimesha Patel, Hatim Ebrahim

**University of Birmingham laboratories:**

Alex Richter, Adrian Shields

## Supplementary Figure

**Supplementary Figure 1.**
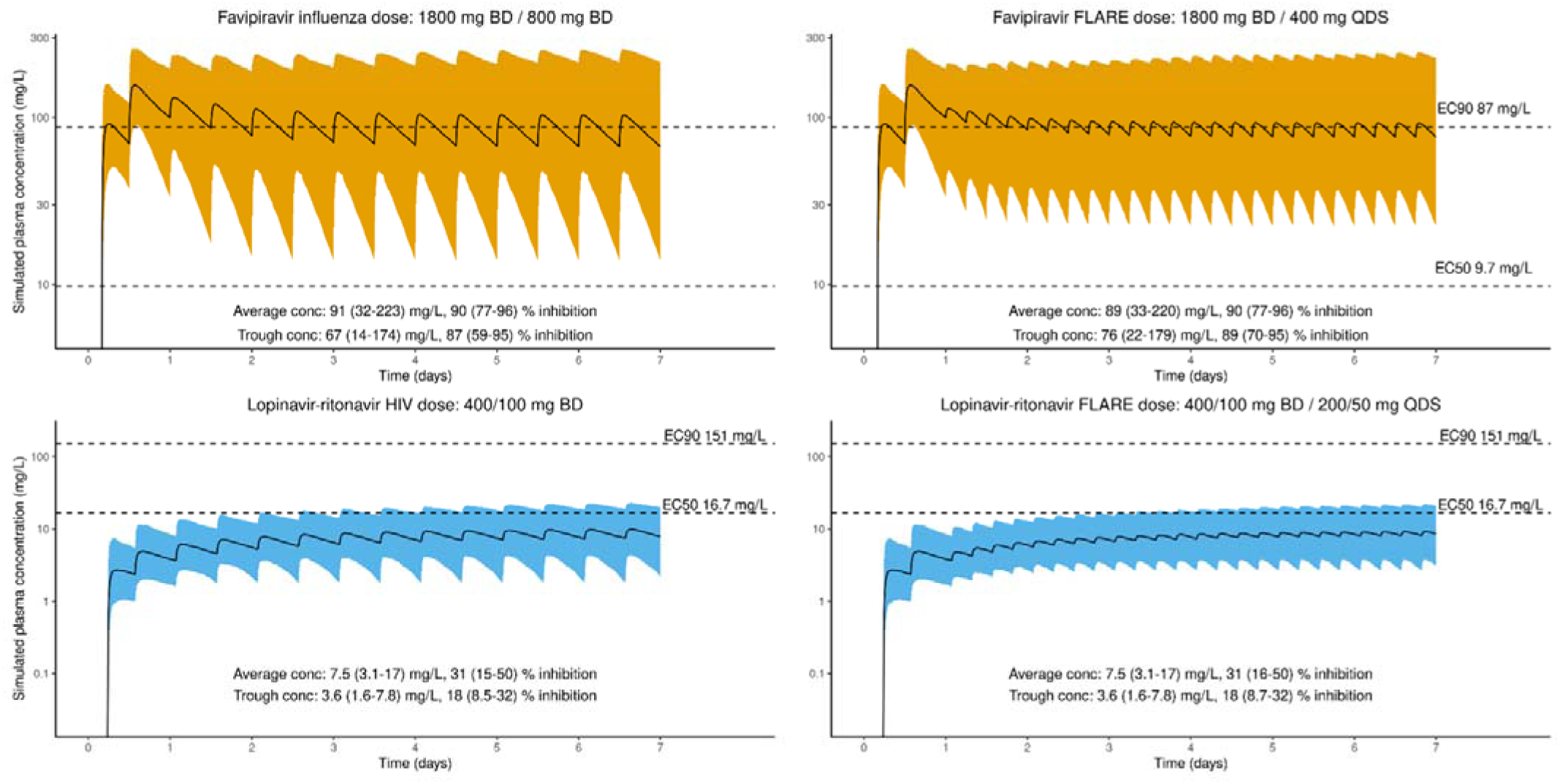
Pharmacometric modelling of predicted plasma concentrations for favipiravir and lopinavir-ritonavir at the doses used in the FLARE trial, presented in relation to the half maximal effective concentration (EC50) and 90% maximal effective concentration (EC90). Simulations are presented for a twice daily (BD) dosing regime and four times daily (QDS) dosing regime.

**Supplementary Figure 2.**
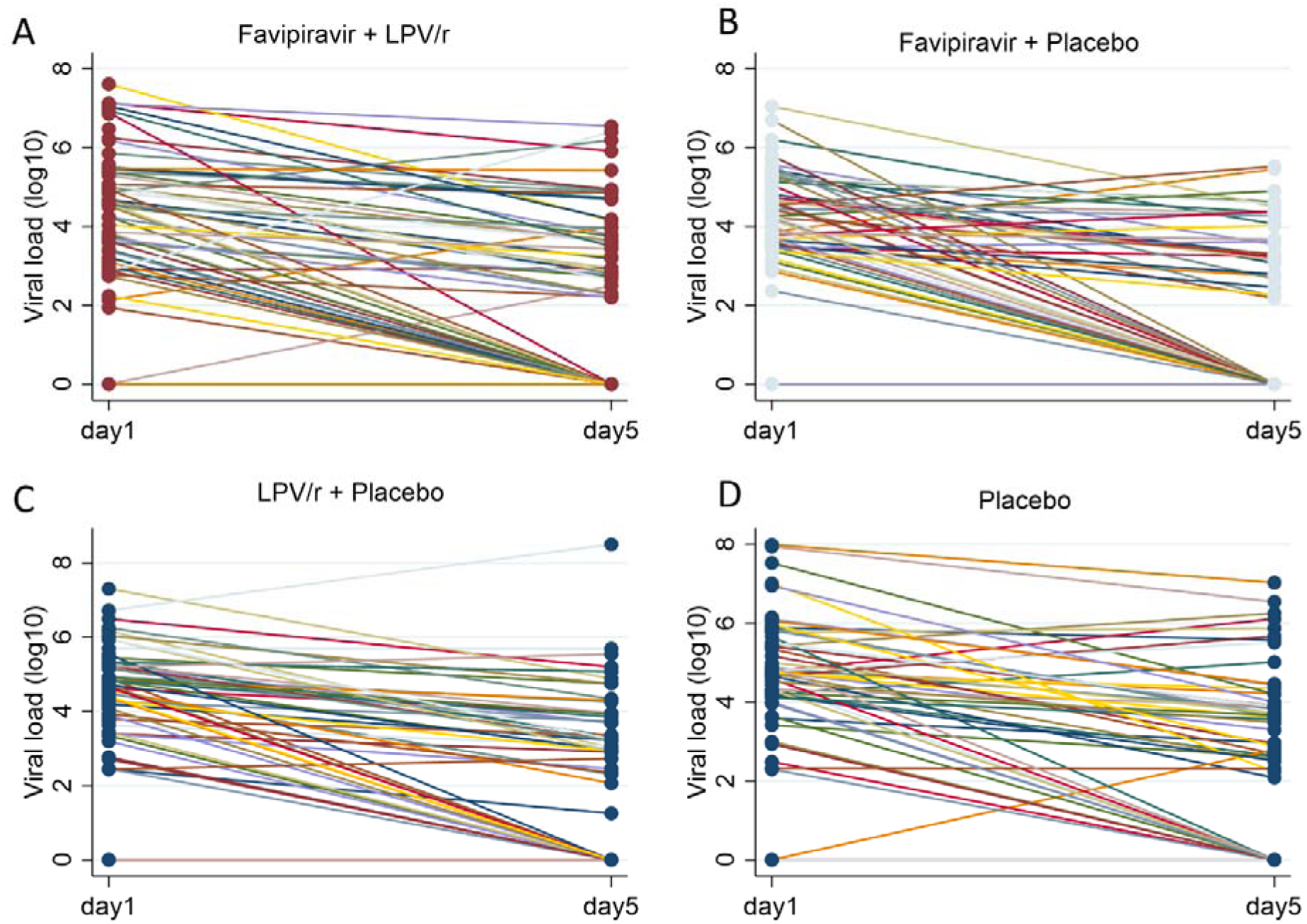
Log_10_ SARS-CoV-2 viral load at baseline (Day 1) and Day 5 presented per participant for (A) favipiravir + lopinavir-ritonavir (LPV/r), (B) favipiravir + placebo, (C) LPV/r + placebo and (D) placebo only.

**Supplementary Figure 3.**
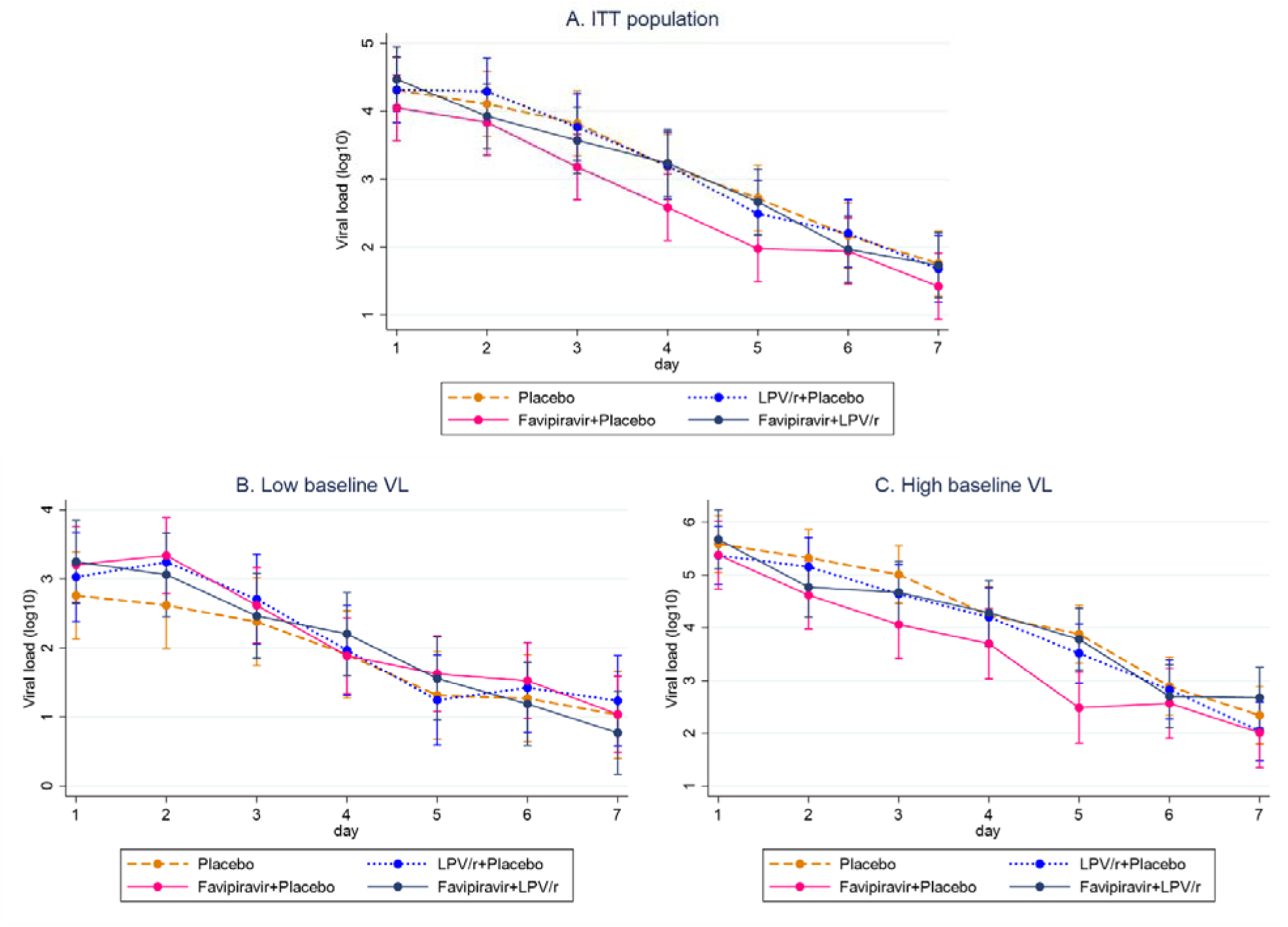
Mean log_10_ SARS-CoV-2 viral load per treatment arm on each day of treatment in (A) the entire cohort, (B) participants with baseline viral load below or equal to the median level and (C) participants with baseline viral load above the median level.

**Supplementary Figure 4.**
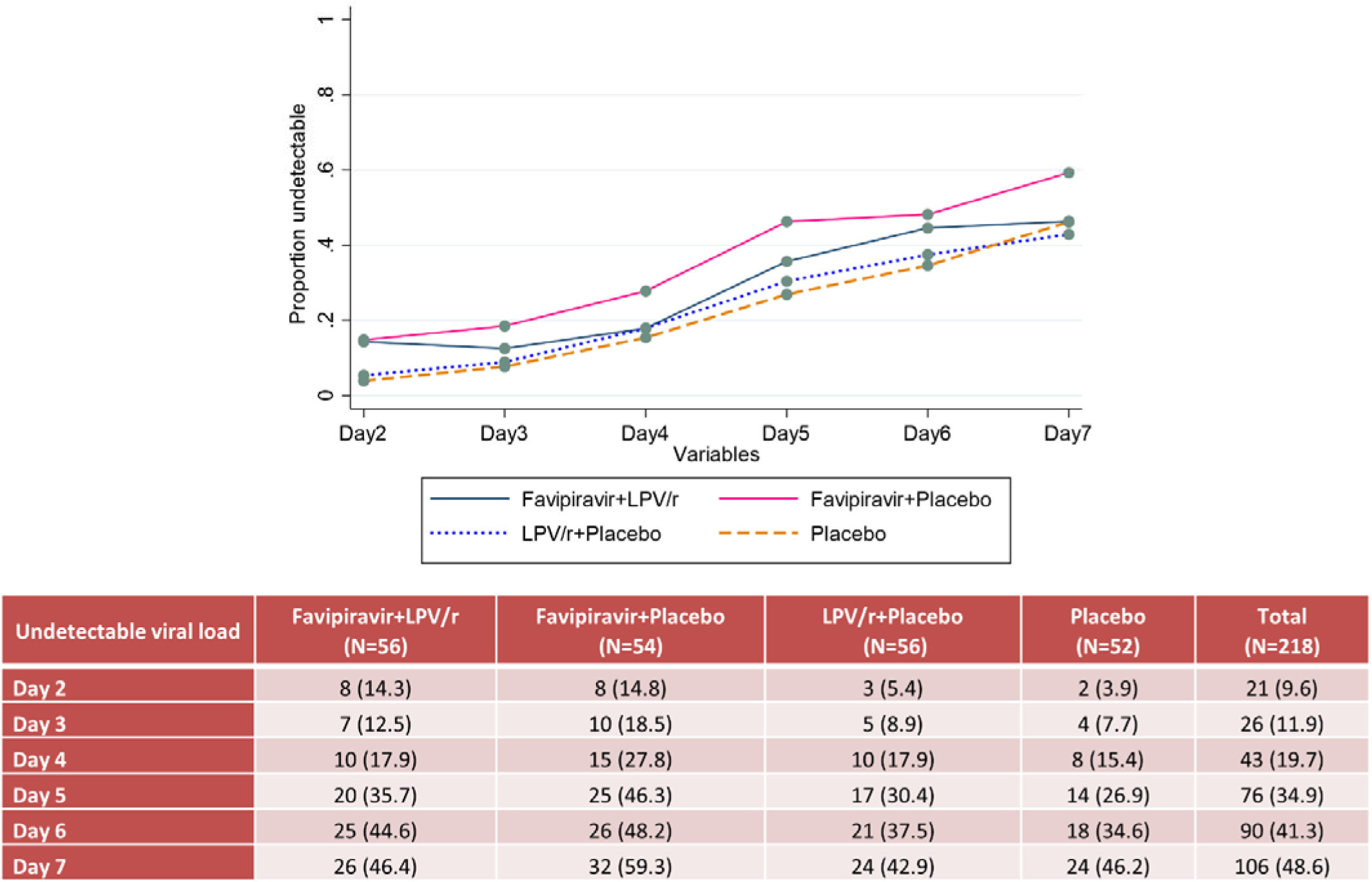
Proportion of participants with detectable viral load at baseline who had undetectable viral load (Ct≥40) on each subsequent day of treatment, per treatment arm. Underlying data are presented in the accompanying table.

**Supplementary Figure 5.**
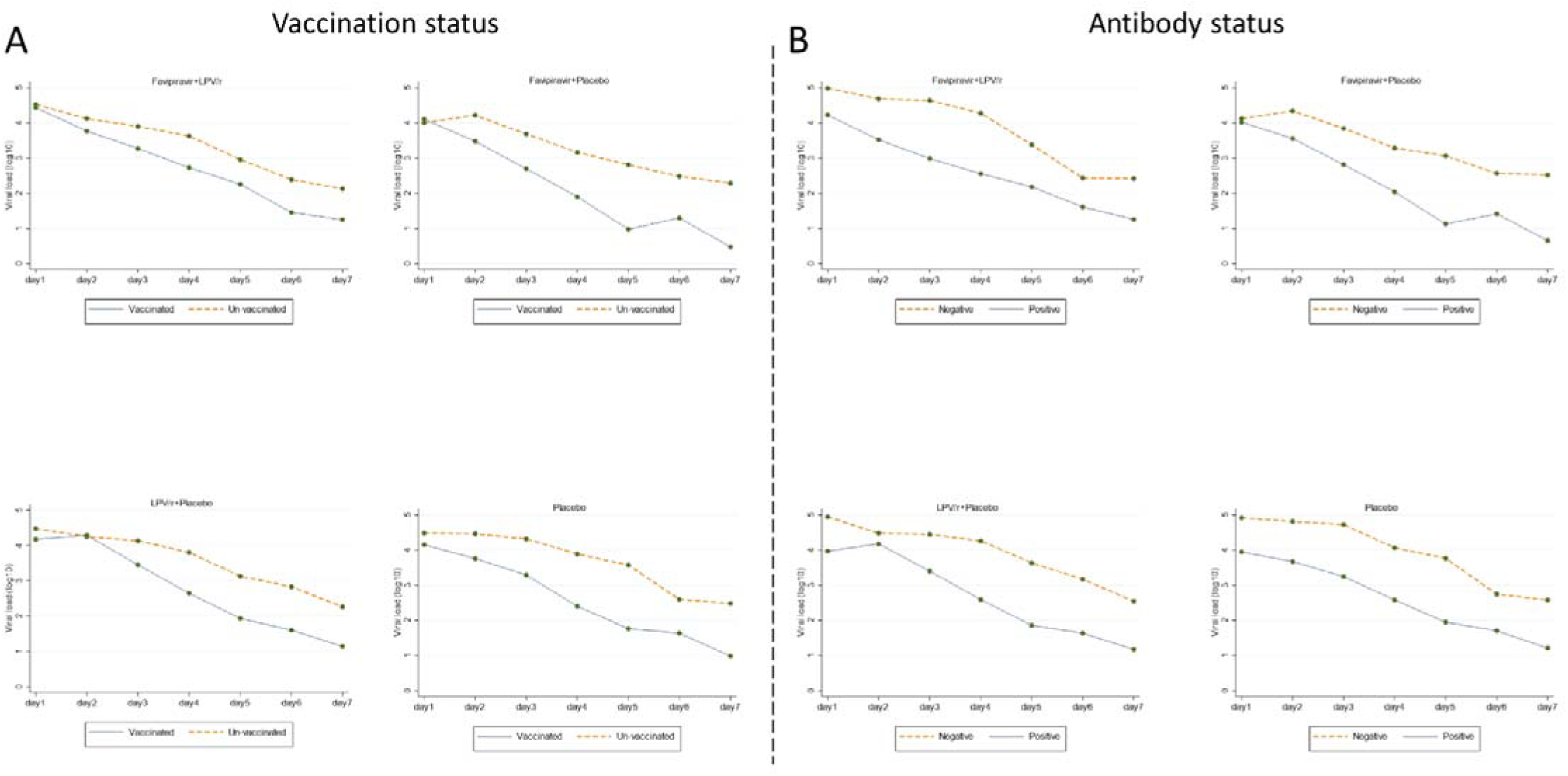
Mean log_10_ SARS-CoV-2 viral load per treatment arm on each day of treatment and per study arm presented (A) according to vaccination status and (B) according to baseline antibody status.

**Supplementary Figure 6.**
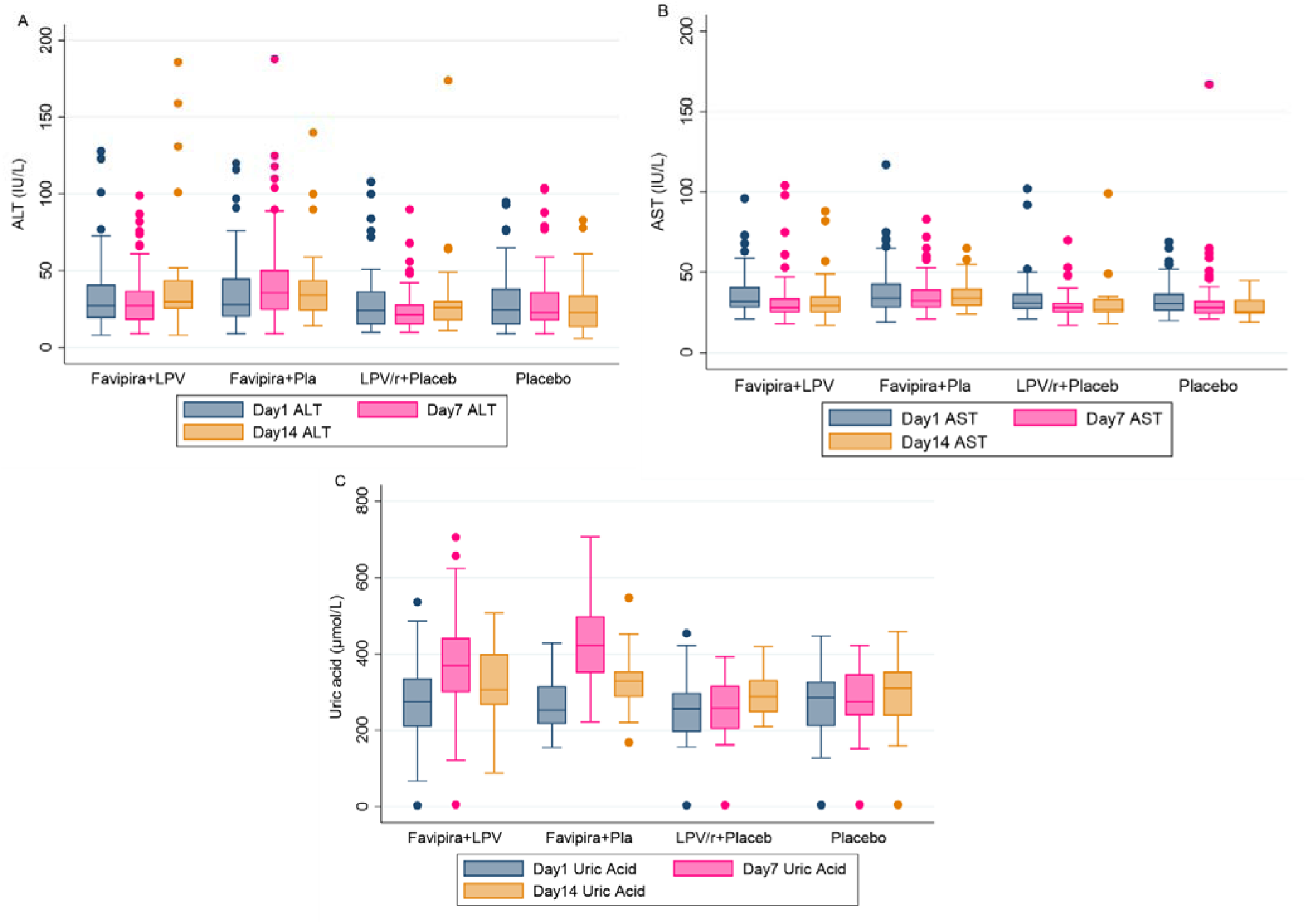
(A) Serum alanine aminotransferase (ALT) concentration, (B) serum aspartate aminotransferase (AST) concentration and (C) serum uric acid concentration at Day 1, Day 7 and Day 14 according to treatment arm. Blood tests were usually only taken at Day 14 if abnormal at Day 7.

## References

1. Gupta A, Gonzalez-Rojas Y, Juarez E, Crespo Casal M, Moya J, Falci DR, et al. Early Treatment for Covid-19 with SARS-CoV-2 Neutralizing Antibody Sotrovimab. N Engl J Med 2021,385:1941–1950.

2. Jayk Bernal A, Gomes da Silva MM, Musungaie DB, Kovalchuk E, Gonzalez A, Delos Reyes V, et al. Molnupiravir for Oral Treatment of Covid-19 in Nonhospitalized Patients. N Engl J Med 2021.

3. Weinreich DM, Sivapalasingam S, Norton T, Ali S, Gao H, Bhore R, et al. REGN-COV2, a Neutralizing Antibody Cocktail, in Outpatients with Covid-19. N Engl J Med 2021,384:238–251.

4. Owen DR, Allerton CMN, Anderson AS, Aschenbrenner L, Avery M, Berritt S, et al. An oral SARS-CoV-2 M(pro) inhibitor clinical candidate for the treatment of COVID-19. Science 2021,374:1586–1593.

5. Syed AM, Ciling A, Khalid MM, Sreekumar B, Chen PY, Kumar GR, et al. Omicron mutations enhance infectivity and reduce antibody neutralization of SARS-CoV-2 virus-like particles. medRxiv 2022.

6. Sanders JM, Monogue ML, Jodlowski TZ, Cutrell JB. Pharmacologic Treatments for Coronavirus Disease 2019 (COVID-19): A Review. JAMA 2020,323:1824–1836.

7. Chu CM, Cheng VC, Hung IF, Wong MM, Chan KH, Chan KS, et al. Role of lopinavir/ritonavir in the treatment of SARS: initial virological and clinical findings. Thorax 2004,59:252–256.

8. Chan KS, Lai ST, Chu CM, Tsui E, Tam CY, Wong MM, et al. Treatment of severe acute respiratory syndrome with lopinavir/ritonavir: a multicentre retrospective matched cohort study. Hong Kong Med J 2003,9:399–406.

9. Park SY, Lee JS, Son JS, Ko JH, Peck KR, Jung Y, et al. Post-exposure prophylaxis for Middle East respiratory syndrome in healthcare workers. J Hosp Infect 2019,101:42–46.

10. Arshad U, Pertinez H, Box H, Tatham L, Rajoli RKR, Curley P, et al. Prioritization of Anti-SARS-Cov-2 Drug Repurposing Opportunities Based on Plasma and Target Site Concentrations Derived from their Established Human Pharmacokinetics. Clin Pharmacol Ther 2020,108:775–790.

11. Kaptein SJF, Jacobs S, Langendries L, Seldeslachts L, Ter Horst S, Liesenborghs L, et al. Favipiravir at high doses has potent antiviral activity in SARS-CoV-2-infected hamsters, whereas hydroxychloroquine lacks activity. Proc Natl Acad Sci U S A 2020,117:26955–26965.

12. Cai Q, Yang M, Liu D, Chen J, Shu D, Xia J, et al. Experimental Treatment with Favipiravir for COVID-19: An Open-Label Control Study. Engineering (Beijing) 2020,6:1192–1198.

13. Ivashchenko AA, Dmitriev KA, Vostokova NV, Azarova VN, Blinow AA, Egorova AN, et al. AVIFAVIR for Treatment of Patients With Moderate Coronavirus Disease 2019 (COVID-19): Interim Results of a Phase II/III Multicenter Randomized Clinical Trial. Clin Infect Dis 2021,73:531–534.

14. Holubar A SA, Purington N, Hedlin H, Bunning B, Walter KS, Bonilla H, Boumis A, Chen M, Clinton K, Dewhurst L, Epstein C, Jagannathan P, Kaszynski RH, Panu L, Parsonnet J, Ponder EL, Quintero O, Sefton E, Singh U, Soberanis L, Truong H, Andrews JR, Desai M, Khosla C, Maldonado Y. Favipiravir for treatment of outpatients with asymptomatic or uncomplicated COVID-19: a double-blind randomized, placebo-controlled, phase 2 trial. medRxiv 2021.11.22.21266690.

15. Ader F, Peiffer-Smadja N, Poissy J, Bouscambert-Duchamp M, Belhadi D, Diallo A, et al. An open-label randomized controlled trial of the effect of lopinavir/ritonavir, lopinavir/ritonavir plus IFN-β-1a and hydroxychloroquine in hospitalized patients with COVID-19. Clin Microbiol Infect 2021,27:1826–1837.

16. Lopinavir-ritonavir in patients admitted to hospital with COVID-19 (RECOVERY): a randomised, controlled, open-label, platform trial. Lancet 2020,396:1345–1352.

17. Gonçalves A, Bertrand J, Ke R, Comets E, de Lamballerie X, Malvy D, et al. Timing of Antiviral Treatment Initiation is Critical to Reduce SARS-CoV-2 Viral Load. CPT Pharmacometrics Syst Pharmacol 2020,9:509–514.

18. Gastine S, Pang J, Boshier FAT, Carter SJ, Lonsdale DO, Cortina-Borja M, et al. Systematic Review and Patient-Level Meta-Analysis of SARS-CoV-2 Viral Dynamics to Model Response to Antiviral Therapies. Clin Pharmacol Ther 2021,110:321–333.

19. Brown LK, Freemantle N, Breuer J, Dehbi HM, Chowdhury K, Jones G, et al. Early antiviral treatment in outpatients with COVID-19 (FLARE): a structured summary of a study protocol for a randomised controlled trial. Trials 2021; 22:193

20. Cook AM, Faustini SE, Williams LJ, Cunningham AF, Drayson MT, Shields AM, et al. Validation of a combined ELISA to detect IgG, IgA and IgM antibody responses to SARS-CoV-2 in mild or moderate non-hospitalised patients. J Immunol Methods 2021,494:113046.

21. Fischer WA, 2nd, Eron JJ, Jr., Holman W, Cohen MS, Fang L, Szewczyk LJ, et al. A Phase 2a clinical trial of Molnupiravir in patients with COVID-19 shows accelerated SARS-CoV-2 RNA clearance and elimination of infectious virus. Sci Transl Med 2021:eabl7430.

22. Nguyen TH, Guedj J, Anglaret X, Laouenan C, Madelain V, Taburet AM, et al. Favipiravir pharmacokinetics in Ebola-Infected patients of the JIKI trial reveals concentrations lower than targeted. PLoS Negl Trop Dis 2017,11:e0005389.

23. Wang Y, Zhong W, Salam A, Tarning J, Zhan Q, Huang JA, et al. Phase 2a, open-label, dose-escalating, multi-center pharmacokinetic study of favipiravir (T-705) in combination with oseltamivir in patients with severe influenza. EBioMedicine 2020,62:103125.

24. Pertinez H, Rajoli RKR, Khoo SH, Owen A. Pharmacokinetic modelling to estimate intracellular favipiravir ribofuranosyl-5’-triphosphate exposure to support posology for SARS-CoV-2. J Antimicrob Chemother 2021,76:2121–2128.

25. Shinkai M, Tsushima K, Tanaka S, Hagiwara E, Tarumoto N, Kawada I, et al. Efficacy and Safety of Favipiravir in Moderate COVID-19 Pneumonia Patients without Oxygen Therapy: A Randomized, Phase III Clinical Trial. Infect Dis Ther 2021,10:2489–2509.

26. Ruzhentsova TA, Oseshnyuk RA, Soluyanova TN, Dmitrikova EP, Mustafaev DM, Pokrovskiy KA, et al. Phase 3 trial of coronavir (favipiravir) in patients with mild to moderate COVID-19. Am J Transl Res 2021,13:12575–12587.

27. Udwadia ZF, Singh P, Barkate H, Patil S, Rangwala S, Pendse A, et al. Efficacy and safety of favipiravir, an oral RNA-dependent RNA polymerase inhibitor, in mild-to-moderate COVID-19: A randomized, comparative, open-label, multicenter, phase 3 clinical trial. Int J Infect Dis 2021,103:62–71.

28. Doi Y, Hibino M, Hase R, Yamamoto M, Kasamatsu Y, Hirose M, et al. A Prospective, Randomized, Open-Label Trial of Early versus Late Favipiravir Therapy in Hospitalized Patients with COVID-19. Antimicrob Agents Chemother 2020,64.

29. Khamis F, Al Naabi H, Al Lawati A, Ambusaidi Z, Al Sharji M, Al Barwani U, et al. Randomized controlled open label trial on the use of favipiravir combined with inhaled interferon beta-1b in hospitalized patients with moderate to severe COVID-19 pneumonia. Int J Infect Dis 2021,102:538–543.

30. Solaymani-Dodaran M, Ghanei M, Bagheri M, Qazvini A, Vahedi E, Hassan Saadat S, et al. Safety and efficacy of Favipiravir in moderate to severe SARS-CoV-2 pneumonia. Int Immunopharmacol 2021,95:107522.

31. Chuah CH, Chow TS, Hor CP, Cheng JT, Ker HB, Lee HG, et al. Efficacy of Early Treatment with Favipiravir on Disease Progression among High Risk COVID-19 Patients: A Randomized, Open-Label Clinical Trial. Clin Infect Dis 2021.

